# Development of Electrospun Nanofibrous Filters for Controlling Coronavirus Aerosols

**DOI:** 10.1101/2020.12.30.20249046

**Authors:** Haihuan Wang, Hongchen Shen, Zhe Zhou, Mengyang Zhang, Minghao Han, David P. Durkin, Danmeng Shuai, Yun Shen

## Abstract

Airborne transmission of SARS-CoV-2 plays a critical role in spreading COVID-19. To protect public health, we designed and fabricated electrospun nanofibrous air filters that hold promise for applications in personal protective equipment and indoor environment. Due to ultrafine nanofibers (∼300 nm), the electrospun air filters had a much smaller pore size compared to the surgical mask and cloth masks (a couple of microns versus tens to hundreds of microns). A coronavirus strain was used to generate aerosols for filtration efficiency tests, which can better represent SARS-CoV-2 than other agents used for aerosol generation in previous studies. The electrospun air filters showed excellent performance by capturing up to 99.9% of coronavirus aerosols, which outperformed many commercial face masks. In addition, since NaCl aerosols have been widely used in filtration tests, we compared the filtration efficiency obtained from the coronavirus aerosols and the NaCl aerosols. The NaCl aerosols were demonstrated as an eligible surrogate for the coronavirus aerosols in the filtration tests, when air filters and face masks with diverse pore sizes, morphologies, and efficiencies were used. Our work paves a new avenue for advancing air filtration by developing electrospun nanofibrous air filters for controlling SARS-CoV-2 airborne transmission. Moreover, the removal efficiency of the NaCl aerosols can be reasonably translated into understanding how air filters capture the coronavirus aerosols.

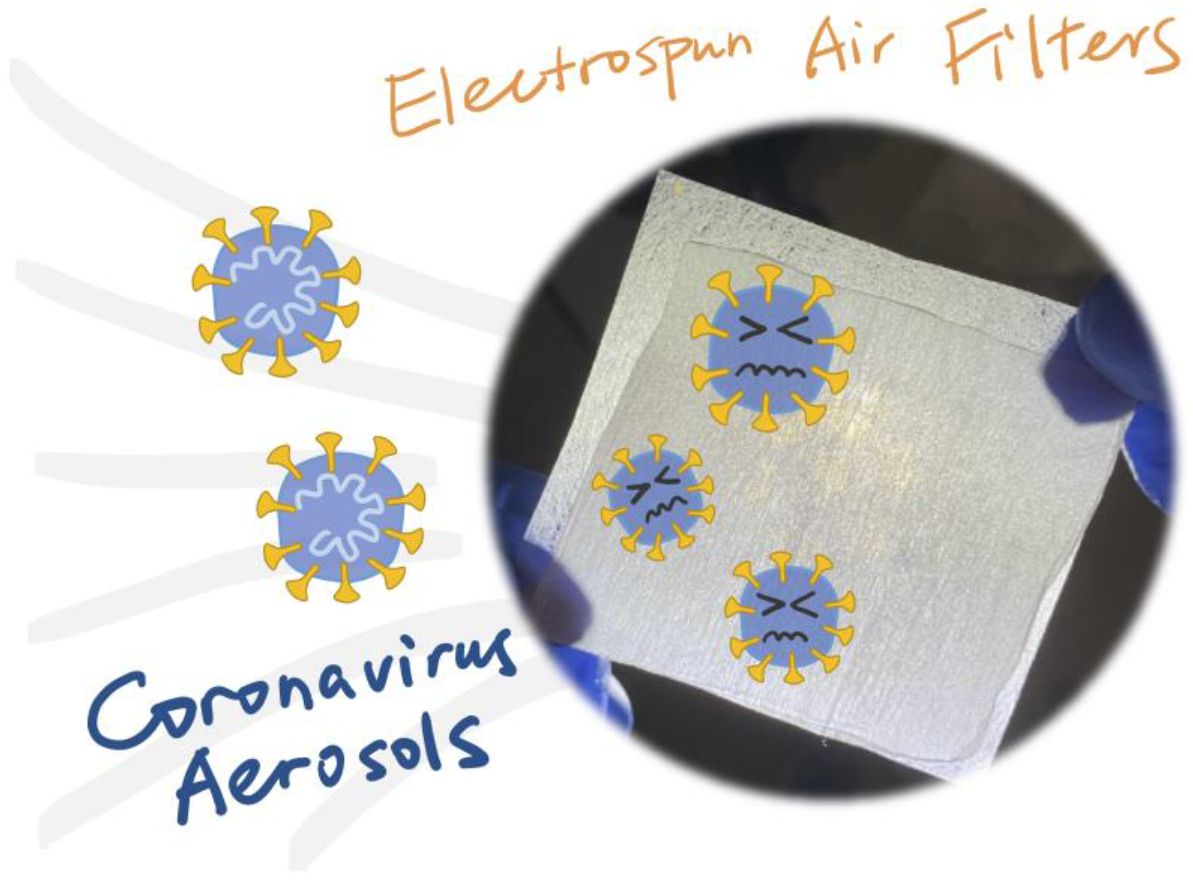
Table of Contents

## Introduction

The ongoing COVID-19 pandemic has raised serious public health concerns. More evidences have highlighted the importance of the airborne transmission of SARS-CoV-2, the causative agent of COVID-19.^12^ Aerosols (< 5 μm) can suspend in the air for a long duration, accumulate in a closed environment, remain infectious (the half-life of SARS-CoV-2 in aerosols is 1.1 h),^3^ and thus involve in the long-range transmission of airborne diseases. Therefore, there is an urgent need to control the airborne transmission of SARS-CoV-2.

One accepted strategy to control the airborne transmission of SARS-CoV-2 is to wear face masks and respirators as personal protective equipment (PPE). However, cloth masks do not always have satisfactory aerosol removal efficiency, droplet repulsion, and/or breathability.^4^ SARS-CoV-2 in the indoor environment can also potentially spread through heating, ventilation, and air conditioning (HVAC) systems,^5^ and effective air filtration is promising to control the spread of airborne diseases. However, most air filters used in residential, commercial, and industrial buildings (minimum efficiency reporting value (MERV) <13) other than high-efficiency particulate air (HEPA) filters used in healthcare facilities only capture larger particles like dusts, mold spores, or bacteria but not airborne viruses.^6^ Nanotechnology holds promise for developing effective, scalable, and affordable air filters for both mask/respirator and HVAC system applications.

Electrospinning has emerged as a new technology to synthesize non-woven nanofibrous membranes that are ideal for air filtration.^7-11^ During electrospinning, a polymer solution is ejected into a strong electric field to form fine nanofibers. Electrospun air filters have a reduced pore size (submicrometers to several micrometers) than conventional filters, which enable effective capture of small airborne particles. The large porosity of electrospun air filters can also reduce air pressure drop in filtration as well as enhance the breathability of masks/respirators.^12^ More importantly, electrospinning is operated under a strong electric field (i.e., 1-5 kV cm^-1^), and it produces filters with retained surface and volume charges that can last for weeks or even months. The presence of the retained charges can significantly promote aerosol capture through electrostatic attraction.^13^ Electrospun nanofibrous membranes have shown excellent performance for removing aerosols generated from polystyrene beads, NaCl, and bacteria (e.g., *Staphylococcus epidermidis, Staphylococcus aureus*).^12, 14-18^

To date, many studies have evaluated the filtration efficiency of air filters and face masks by using surrogate aerosols instead of coronavirus aerosols.^12, 14-20^ However, it remains elusive whether the reported filtration performance can be used to understand coronavirus control because of distinct properties between the surrogate and coronavirus. Moreover, developing electrospun air filters for capturing viral aerosols is still at its nascent stage and no report is available yet. In this work, we systematically compared the filtration efficiency of coronavirus and NaCl aerosols for a broad spectrum of electrospun air filters and face masks, and concluded that NaCl is an eligible surrogate for coronavirus during aerosol filtration tests for the first time. Murine hepatitis virus A59 (MHV-A59), a β-coronavirus in the same family as SARS-CoV-2, was selected for aerosol generation and filtration. In addition, we developed advanced electrospun air filters for capturing coronavirus aerosols that outperformed many commercially available face masks.

## Materials and Methods

### Fabrication of electrospun air filters

We used a customized electrospinning system^21^ to prepare electrospun air filters, by electrospinning a polyvinylidene fluoride (PVDF, 15 wt %, Arkema KYNAR® 761) solution onto a layer of polypropylene fabrics (PP, dissembled from VWR® basic protection face mask, 414004-680). The feeding rate of the working solution, electric field, and electrospinning duration was at 0.6 mL h^-1^, 1 kV cm^-1^, and 20 or 30 min, respectively. The fabricated electrospun air filters were designated as PVDF_20_ and PVDF_30_, respectively. To increase the binding between coronavirus and air filters by electrostatic attraction and promote virus removal efficiency, a positive or negative charged polyelectrolyte, i.e., poly(ethyleneimine) (PEI) or poly(vinylphosphonic acid) (PVPA), was coated onto PVDF_20_, and the fabricated filters were denoted as PVDF_20_/PEI and PVDF_20_/PVPA, respectively. Details of electrospinning, PVPA synthesis, and coating were described in the Supporting Information (SI).

### Characterization of electrospun air filters and face masks

A three-layer non-woven surgical mask, a three-layer woven cotton mask, and a one-layer woven polyester neck gaiter were selected as representative commercial face masks because of their wide availability, extensive use, and distinct properties. The fiber diameter and elemental mapping of the electrospun air filters and the face masks were determined by scanning electron microscopy with energy-dispersive X-ray spectroscopy (SEM-EDS), PEI or PVPA coating was characterized by attenuated total reflectance-Fourier transform infrared (ATR-FTIR) spectroscopy, mean flow pore size was examined by a gas liquid porometry method, and pressure drop was determined with a face velocity of 5.3 cm s^-1^. Details are included in the SI.

### Determination of filtration efficiency for coronavirus and NaCl aerosols

The filtration efficiency was evaluated by a customized aerosolization setup (CH Technologies, Inc., **Figure S1**). Briefly, the aerosols were generated by a high output Blaustein Atomizer (BLAM) with four jets, and high-rate air flow (∼1.5 L min^-1^) sheared liquid flow (15 mL h^-1^) of a NaCl solution (0.1 M) or MHV-A59 in phosphate buffered saline (PBS, ∼10^6^ gene copies mL^-1^) into tiny droplets including aerosols. The BLAM produced aerosols with a whole size distribution of 200 nm-3.5 μm, and ∼90% of the aerosols were ranged between 200 to 400 nm based on the number of aerosols. A single pass atomization (no air circulation) mode was applied to ensure the least damage of viruses during aerosolization. A portion of the aerosols with a controlled flow rate of 0.5 standard liter per minute next passed through a fitted air filter/face mask mounted on a stainless-steel holder and an impinger sequentially, and each filtration test (i.e., filter-on) was conducted for 30 min. The rest aerosols were captured by a HEPA filter under vacuum. NaCl or MHV-A59 aerosols that penetrated through the filter/mask were retained by the impinger in ultrapure water or PBS (4 mL), respectively. Control experiments (i.e., filter-off) with the same setup and operational conditions were also conducted, except that no filter was installed in the holder. Each filter-on and filter-off experiment was conducted at least for duplicates, and filtration efficiency, including mean, max, and min values, was calculated from the difference of the amount of the testing agent (NaCl or MHV-A59) in the impinger between filter-off and filter-on experiments over the amount of the testing agent in the impinger in the filter-off experiment. Details of quantifying the testing agent in the impinger and calculating the filtration efficiency are shown in the SI.

## Results and Discussion

### Electrospun air filters had small fiber diameters and pore sizes

Comparing with the commercial face masks, the electrospun PVDF filters fabricated in this study were with much smaller fiber diameters and pore sizes (**Figures 1** and **S2**). Specifically, the fiber diameter of PVDF_20_, PVDF_30_, PVDF_20_/PEI, and PVDF_20_/PVPA ranged 0.2-1.3 μm. Compared to PVDF_20_, PVDF_30_ showed a similar fiber diameter and pore size (t-test, both p>0.05), but its increased thickness induced a slightly higher pressure drop (0.23±0.00 versus 0.20±0.00 inches of water column (inch wc)). Both elemental mapping and FTIR characterization indicated the successful coating of PEI or PVPA onto the electrospun PVDF air filters (**Figures S3** and **S4**). The fiber diameter of PVDF_20_/PEI and PVDF_20_/PVPA did not change (t-test, both p>0.05) in comparison with PVDF_20_. However, PEI and PVPA were observed to block pores in air filters. For PVDF_20_/PVPA, its pores were partially blocked (**Figure S2d**), leading to a smaller pore size than PVDF_20_ (t-test, p=0.006). For PVDF_20_/PEI, some pores especially the small ones were completely blocked (**Figure S2c**), but the mean pore size was comparable with that of PVDF_20_ (t-test, p=0.27). Both PVDF_20_/PEI and PVDF_20_/PVPA showed an increased pressure drop of 1.19± 0.59 and 0.38±0.04 inch wc than PVDF_20_.

**Figure 1.**
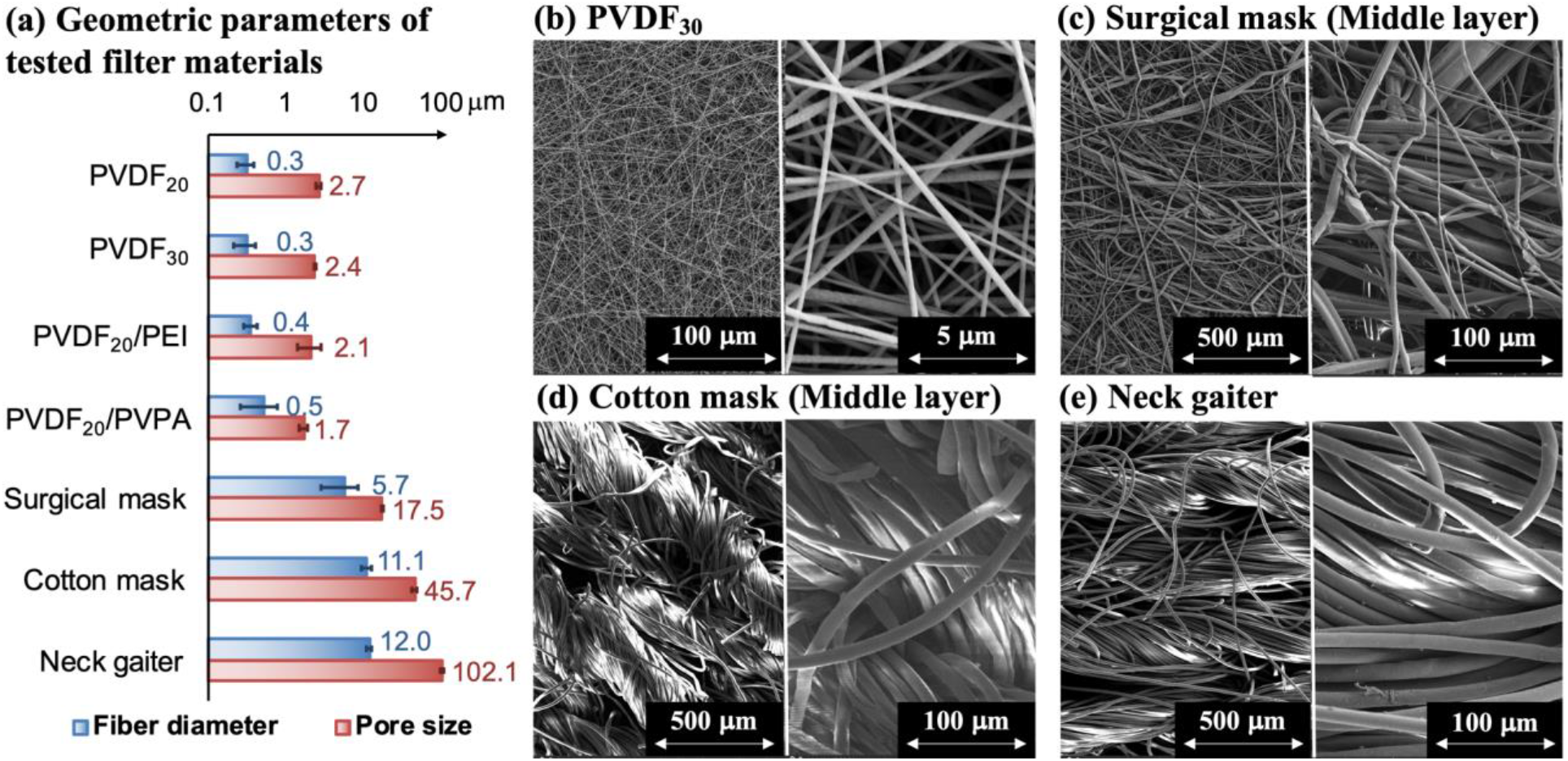
(a) Geometric parameters of tested filter materials, and SEM of (b) PVDF_30_, (c) surgical mask middle layer, (d) cotton mask middle layer, and (e) neck gaiter.

Among the three commercial face masks, the middle layer of the surgical mask, which is the effective filtration medium, showed the smallest fiber diameter of 5.7±2.8 μm, while the neck gaiter showed the largest fiber diameter of 12.0±1.0 μm. A larger fiber diameter corresponded to a larger pore size: all the electrospun filters had a mean pore size ≤ 2.7 μm, all the commercial masks had the mean pore size ≥17.5 μm, and the largest mean pore size of 102.1±4.4 μm was observed for the neck gaiter. The positive correlation between the filter fiber diameter and the pore size agreed with previous studies.^22^ The filters with a larger fiber diameter and pore size showed a lower filtration efficiency for airborne particles.^23^ Therefore, we expect that electrospun air filters would outperform the commercial face masks for removing airborne coronavirus particles. Due to a large pore size, the surgical mask, cotton mask, and neck gaiter has a low pressure drop of 0.06±0.01, 0.06±0.01, and 0.01±0.00 inch wc, respectively.

### Electrospun air filters showed a high filtration efficiency for removing coronavirus aerosols

All the electrospun filters had an average filtration efficiency ≥95.7%, while the commercial face masks showed an average filtration efficiency of 44.9% for the neck gaiter, 73.3% for the cotton mask, and 98.2% for the surgical mask, respectively (**Figure 2**). The aerosol removal efficiency indeed increased with the decrease of the mask pore size.^22, 23^ Increase of electrospinning duration and thickness of the air filters enhanced coronavirus aerosol removal (99.9% and 99.1% for PVDF_30_ and PVDF_20_, respectively), though there was a marginal decrease of the pore size of PVDF_30_ compared to PVDF_20_. Polyelectrolyte coating did not promote electrospun air filters for removing coronavirus aerosols, and the average filtration efficiency for PVDF_20_/PEI and PVDF_20_/PVPA was 99.1% and 95.7%, respectively. We speculate that coronavirus aerosol capture was dominated by interception, impaction, or diffusion, instead of the long-range force of electrostatic attraction, since the size of the aerosols and electrospun filter pores are close. The fact that PVPA coating reduced the aerosol removal efficiency may indicate coronavirus aerosols are negatively charged under the experimental condition, and electrostatic repulsion might play a role in filtration. The PP fabric for supporting the electrospun membrane removed a very limited amount of aerosols (0-30%), suggesting that the electrospun layer determines the performance of virus capture. Details are included in the SI.

**Figure 2.**
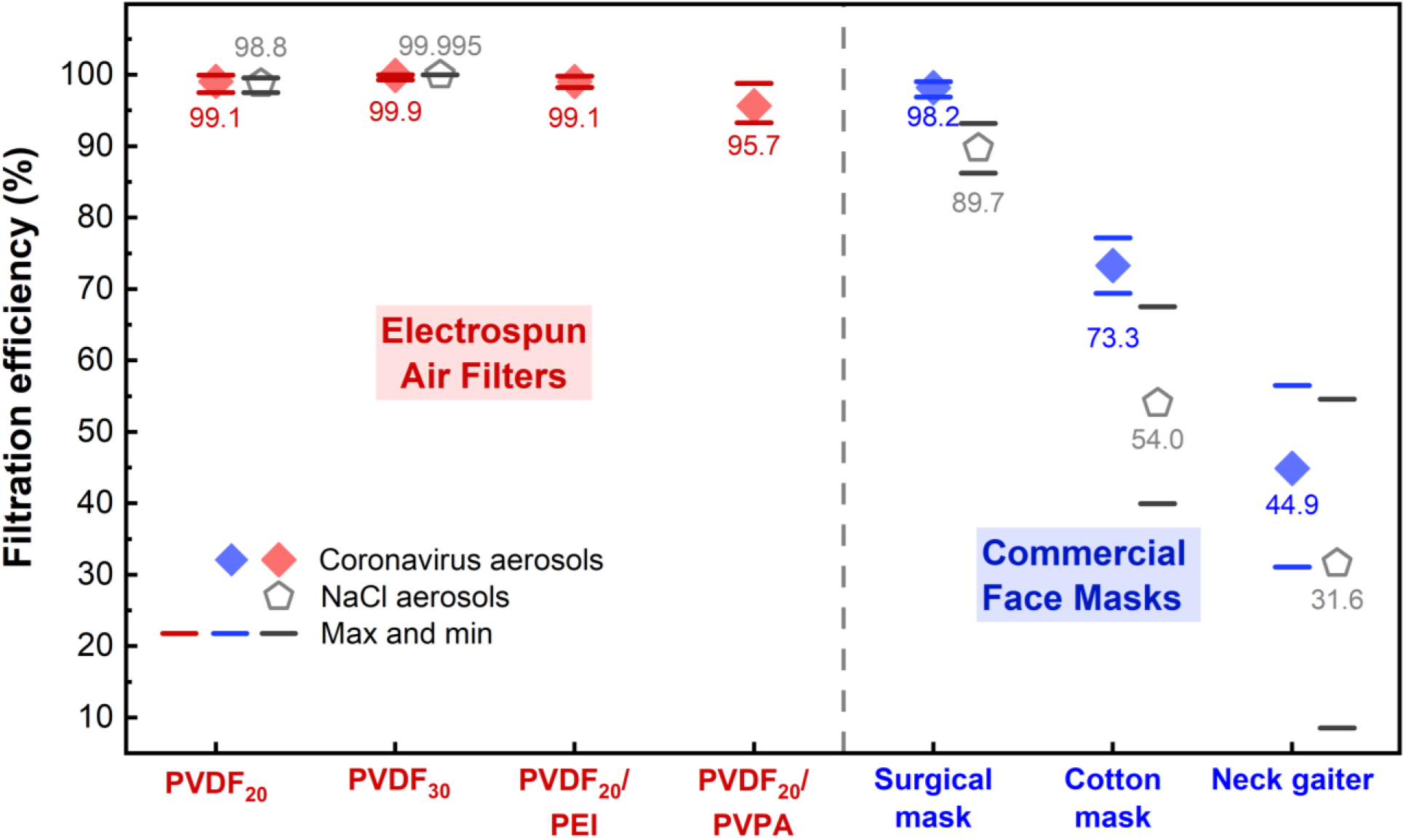
Aerosol filtration efficiency of electrospun air filters and commercial face masks. Aerosols generated from coronavirus (MHV-A59) and NaCl were used for tests. Red and blue diamonds represent the average filtration efficiency of MHV-A59 aerosols by the electrospun air filters and the commercial face masks, respectively. Gray pentagon represents the average filtration efficiency of NaCl aerosols. Red, blue, and gray bars represent max and min values of the filtration efficiency in replicates.

To the best of our knowledge, our study is the first to evaluate the filtration efficiency of the air filters/face masks by using aerosols of a coronavirus rather than surrogates. A previous study used bacteriophage MS2 to produce aerosols, and it revealed that the surgical mask removed 98.3-100% of the viral aerosols.^19^ However, MS2 is a non-enveloped virus with an average size of 27 nm, which is very different from SARS-CoV-2 with an envelope structure and a virion size of 50-200 nm.^24^ In addition, that study only tested the filtration of aerosols with a size of several microns (2.6 and 6.0 μm), while the filtration efficiency of smaller aerosols is still unknown.^19^ Aerosol particles with a size of 100-500 nm can suspend in air for extended duration and they are most penetrating for mechanical filters,^25, 26^ but filtration of viral aerosols with those sizes has not been studied yet. The surgical mask was reported to remove 76-90% of silica or NaCl aerosols with a size around 300 nm,^4, 27^ and the efficiency was lower than that in our study for capturing coronavirus aerosols with a dominant size between 200 to 400 nm. The difference between the filtration efficiency could be caused by distinct physicochemical properties (e.g., size, geometry, surface charge, and elasticity) of viral and inorganic particles/aerosols. Compared to the aerosols produced from inert agents,^4, 19, 27^ MHV-A59 aerosols used in our study can best represent SARS-CoV-2 aerosols.

### NaCl aerosols served as an eligible surrogate for evaluating the filtration efficiency of coronavirus aerosols

The filtration efficiencies for removing coronavirus and NaCl aerosols were compared for PVDF_20_, PVDF_30_, and the commercial face masks (**Figure 2**). For electrospun air filters, the cotton mask, and the neck gaiter, using MHV-A59 or NaCl aerosols did not lead to significant difference in filtration efficiency (t-test, all p>0.05). The filtration efficiency of NaCl aerosols for the surgical mask was significantly lower than that of MHV-A59 aerosols (t-test, p=0.00001). Therefore, NaCl aerosols can serve as a good surrogate for coronavirus aerosols in evaluating the filtration efficiency for different air filters and face masks, because the removal efficiency for NaCl aerosols was lower or equivalent to that for coronavirus aerosols. Since tests with pathogenic viral aerosols require high biosafety levels, NaCl aerosols are more easily to handle and they have been frequently used for filtration tests.^4, 28-30^ NaCl and bacteriophage phiX174 aerosols were used to test the filtration efficiency of the N95 respirator and the surgical mask, and a similar or lower filtration efficiency was also observed for NaCl aerosols compared to phiX174 aerosols.^20^ However, phiX174 is a non-enveloped virus with a virion size of 27 nm, which may not well represent SARS-CoVo-2. In addition, both the N95 respirator and the surgical mask had relatively high filtration performance, regardless of the type of aerosols used.^20^ The conclusion that NaCl aerosols can act as a surrogate for viral aerosols based on the tests of high-performance filters may not be valid for low-performance filters. Our study has addressed this concern by demonstrating NaCl aerosols are a valid surrogate for coronavirus aerosols for both high- and low-performance filters.

### Implication

Our study is the first to demonstrate that electrospun air filters hold promise for providing efficient protection against airborne coronavirus particles. Cloth masks (e.g., cotton mask and neck gaiter) are most commonly used during the COVID-19 pandemic, compared to the surgical mask and N95 respirator;^31^ but our study along with previous research revealed the a low filtration efficiency of the cloth masks.^19, 27^ Therefore, wearing the surgical mask or other masks with an equal or higher filtration efficiency should be encouraged to protect people from SARS-CoV-2 airborne transmission and infection. However, at the beginning of the pandemic, the shortage of the surgical mask and the N95 respirator was frequently reported, and it calls for an urgent need to advance air filter development for PPE. Meanwhile, it is highly desirable to develop innovative and efficient air filters for the HVAC system that can prevent the long-range transmission and accumulation of coronavirus aerosols. Electrospinning is an economically feasible and industrially viable technology to fabricate highly efficient air filters,^32, 33^ and our study underscores its great potential for controlling the spread of coronavirus aerosols. More attractively, small-scale electrospinning apparatuses can provide a rapid response to the pandemic and produce highly tailorable filters with desired performance for individuals and small communities.^34, 35^ By integrating with additive manufacturing (i.e., 3D printing),^36^ it is easy to design and fabricate customized masks, respirators, and HVAC air filters based on the needs onsite.

Our study has also validated the eligibility of NaCl as a surrogate of coronavirus in aerosol filtration tests, by comparing the filtration efficiency for a broad spectrum of air filters and face masks with different pore sizes and ranges of efficiency. Research outcome can be translated into understanding how to leverage filtration for controlling coronavirus airborne transmission when NaCl aerosols were tested. Precautions should be paid to using our conclusion for interpreting other research results, because we do not know yet how the charge and size of coronavirus aerosols, the charge of filters, and other experimental conditions (e.g., flow rate of aerosols) impact the surrogate validity of NaCl for coronavirus. Future work should explore these items.

## ASSOCIATED CONTENT

The Supporting Information is available free of charge on the ACS Publications website at DOI: XXX.

Electrospinning; PVPA synthesis; material characterization; polyelectrolyte coating; MHV-A59 culturing and RT-qPCR quantification; NaCl quantification; filtration efficiency calculation; filtration efficiency comparison between the electrospun layer and the PP supporting fabric; scheme of air filtration setup; SEM and elemental mapping of electrospun air filters and face masks; ATR-FTIR characterization of polyelectrolyte coated filters **(PDF)**.

## Data Availability

All data are available in the manuscript.

## Author Information

## Acknowledgements

We acknowledge the NSF RAPID grants (CBET-2029411 and CBET-2029330) for supporting our research. We thank The George Washington University (GW) Nanofabrication and Imaging Center for SEM characterizations, GW BSL2+ lab facilities for the bioaerosol study, and United States Naval Academy for ATR-FTIR characterizations. We also thank I. Kienbaum and Kees van der Kamp in APTCO Technologies LLC for performing the pore size analysis for the air filters and face masks.

