## Supporting Information for "Development of Electrospun Nanofibrous Filters for Controlling Coronavirus Aerosols"

#### Supporting Information

Haihuan Wang,[1,2#] Hongchen Shen,[1#] Zhe Zhou,[1#] Mengyang Zhang,[1] Minghao Han,[2] David P. Durkin,[3] Danmeng Shuai,[1*] Yun Shen[2*]

1 Department of Civil and Environmental Engineering, The George Washington University, Washington, DC 20052

2 Department of Chemical and Environmental Engineering, University of California, Riverside, Riverside, CA 92521

3 Department of Chemistry, United States Naval Academy, Annapolis, MD 21402

### Equal Contribution.

* Corresponding Authors:

Danmeng Shuai: Phone: 202-994-0506, Fax: 202-994-0127,, Website: http://materwatersus.weebly.com/

15 Pages, 10 Texts, 4 Figures

**Text S1. Electrospinning of Air Filters**

A homogeneous working solution was first prepared by dissolving 15 wt % of polyvinylidene fluoride (PVDF, Arkema KYNAR® 761) in a mixture of *N*,*N*-dimethylformamide (DMF) and acetone (DMF: acetone = 7:3, v/v). Next, the working solution was used to electrospin a nanofibrous membrane onto a layer of polypropylene fabrics (PP, dissembled from VWR® basic protection face mask, 414004-680) attached on a rotating drum collector (600 rpm), which produced a composite electrospun air filter. The PP fabrics provided mechanical support of the electrospun membrane. The feeding rate of the working solution, electric field, and electrospinning duration was at 0.6 mL $h^{-1}$, 1 kV $cm^{-1}$ (10 kV of voltage and 10 cm between the electrospinning needle and the drum collector), and 20 or 30 min, respectively. The fabricated electrospun air filters were designated as $PVDF_{20}$ and $PVDF_{30}$, respectively.

**Text S2. Synthesis of Poly(vinylphosphonic acid) (PVPA)**

PVPA was synthesized according to the previous literature.[1] 1 g of vinylphosphonic acid (Sigma-Aldrich, 97%), 2.5 mg of 2,2'-azobis(2-methylpropionamidine) dihydrochloride (Sigma-Aldrich, 97%), and 0.5 mL of ultrapure water were added in a Schlenk flask for polymerization. Nitrogen gas was flushed into the flask before the polymerization. The mixture was then heated at 80 °C for 3 h, the resulted viscous solution contains PVPA.

**Text S3. Coating of Electrospun Membranes with Polyelectrolytes**

The electrospun PVDF membrane on the PP fabric was pre-wetted by ethanol and then immersed in a PEI or PVPA solution (5 wt % in an ethanol-water mixture (50/50, w/w)) for 10 min. The composite membrane was finally dried via vacuum filtration and stored in a desiccator.

**Text S4. Characterization of Electrospun Air Filters and Face Masks**

The morphology, fiber diameter, and elemental mapping of the electrospun filters and the face masks were determined by scanning electron microscopy with energy-dispersive X-ray spectroscopy (SEM-EDS, FEI Teneo LV). For each filter or face mask, at least 50-100 fibers from different SEM images were used for fiber diameter analysis. Elemental mapping of C, N, P, and O and attenuated total reflectance-Fourier transform infrared (ATR-FTIR) spectroscopy (iS10 Nicolet Thermo ATR-FTIR) were characterized for $PVDF_{20}$/PEI and $PVDF_{20}$/PVPA. The mean flow pore size of the filters/masks was examined by a gas liquid porometry method (POROLUX™ 100/200/500, shape factor of 0.715, APTCO Technologies LLC, Belgium). For each sample, as least three different pieces were characterized. Pressure drop across the filters/masks was determined with a face velocity of 5.3 cm $s^{-1}$ by an accurate differential pressure gage (Dwyer, Magnehelic®, 2001-HA), and at least duplicates were conducted for each sample.

**Text S5. Culturing and Purification of Murine Hepatitis Virus A59 (MHV-A59)**

MHV-A59 was cultured in HeLa-mCC1a cells according to the literature.[2] HeLa-mCC1a cells were maintained in Dulbecco's modified Eagle's medium (DMEM) supplemented with 10% fetal bovine serum (FBS), 1% Penicillin-Streptomycin (P/S), and 500 µg $mL^{-1}$ of G418 sulfate and incubated at 37 °C/5% $CO_2$ until confluence. DMEM and DMEM supplemented with both FBS and P/S are referred as the serum free medium (SFM) and the complete culture medium, respectively. For MHV-A59 stock propagation and enrichment, confluent HeLa-mCC1a cells were first washed with SFM for three times, infected with MHV-A59, and incubated at 37 °C/5% $CO_2$ until a significant cytopathic effect was observed. Cells were next frozen and thawed for three times, the suspension was centrifuged (1,000 ×g, 15 min) to remove cell debris after a couple of times of sonication. The supernatant was collected and stored at -80 ℃ until use.

**Text S6. RNA Extraction and RT-qPCR Quantification of MHV-A59**

Viral RNA was extracted by a Zymo Quick-RNA Viral Kit (R1035). RT-qPCR was conducted using the TaqMan™ Fast Virus 1-Step Master Mix kit (Thermo Fisher Scientific Inc., 4444432), which combines reverse transcription (RT) reaction with quantitative PCR together. Each 20 μL of reaction mix contained 5 μL of RNA template, 5 μL of Fast Virus Master Mix, 2 μL of forward primer (10 μM), 2 μL of reverse primer (10 μM), 1.25 μL of probe, and 4.75 μL of RNA-free water. The primer was designed in the sequence of ORF5 gene of MHV-A59, which is corresponding to structural protein M. The hydrolysis probe was applied for detection, which consisted of the fluorescein (FAM), i.e., a fluorescent reporter, at the 5' end and the Black Hole Quencher 1 dye (BHQ1), i.e., a quencher, at the 3' end. The absolute genome copy numbers were standardized using a series of 10-fold dilutions ($10^3$-$10^{10}$ copies $mL^{-1}$) of synthetic cDNA oligo (IDT). The RT-qPCR program was run on the QuantStudio (Applied Biosystems) instrument: reverse transcription (52 ℃ for 10 min), reverse transcription inactivation and denaturation initiation (60 ℃ for 20 s), and 45 cycles of amplification (95 ℃ for 15 s and 60 ℃ for 60 s). RNA-free water was used as the negative control.

The sequences of primers for MHV-A59 were GGAACTTCTCGTTGGGCATTATACT and ACCACAAGATTATCATTTTCACAACATA; the sequence of the probe for MHV-A59 was 56-FAM/ACATGCTACGGCTCGTGTAACCGAACTGT/3BHQ_1, and the sequence of the DNA standard for MHV-A59 was

CTTAAGGAATGGAACTTCTCGTTGGGCATTATACTACTCTTTATTACTATCATACTAC AGTTCGGTTACACGAGCCGTAGCATGTTTATTTATGTTGTGAAAATGATAATCTTGT GGTTAATGTGGCC.

**Text S7. Preparation of an MHV-A59 Solution for Aerosolization**

A solution of MHV-A59 in phosphate buffered saline (PBS) was prepared for aerosolization. Briefly, MHV-A59 was cultured (**Text S5**), quantified by reverse-transcription quantitative polymerase chain reaction (RT-qPCR) targeting on the ORF5 gene (**Text S6**), purified by centrifugal ultrafiltration to remove culture media (Nanosep, 300 kDa, Pall Laboratory), and diluted in PBS to a concentration of ~$10^6$ gene copies $mL^{-1}$ for aerosolization.

**Text S8. Quantification of NaCl and MHV-A59 in the Impinger**

NaCl concentration was quantified by ion chromatography (Dionex ICS-1100; Dionex IonPac™ AS18 column) using 10 mM NaOH as eluent and 0.25 mL $min^{-1}$ as the eluent flow rate. The amount of NaCl in the impinger was calculated from its concentration and the volume of solution in the impinger (4 mL). For quantifying MHV-A59, 3.5 mL of collected MHV-A59 solution in the impinger was first concentrated to ~100 μl by centrifugal ultrafiltration (Nanosep, 300 kDa, Pall Laboratory), the RNA of the concentrated solution was next extracted by a Zymo Quick-RNA Viral Kit (R1035) and quantitatively determined by RT-qPCR. The total amount of MHV-A59 in the impinger was calculated by multiplying 1.14 (= 4 mL/3.5 mL) of the value reported by RT-qPCR, with a unit of gene copies.

**Text S9. Filtration Efficiency Evaluation**

$$Mean\ Filtration\ Efficiency = \frac{\frac{\sum_{j=1}^{m} b_j}{m} - \frac{\sum_{i=1}^{n} a_i}{n}}{\frac{\sum_{j=1}^{m} b_j}{m}} \times 100\% \quad \text{(Eq. S1),}$$

$$Max\ Filtration\ Efficiency = \frac{\max(b_1, b_2, \ldots, and\ b_m) - \min(a_1, a_2, \ldots, and\ a_n)}{\max(b_1, b_2, \ldots, and\ b_m)} \times 100\% \quad \text{(Eq. S2),}$$

$$Min\ Filtration\ Efficiency = \frac{\min(b_1, b_2, \ldots, and\ b_m) - \max(a_1, a_2, \ldots, and\ a_n)}{\min(b_1, b_2, \ldots, and\ b_m)} \times 100\% \quad \text{(Eq. S3),}$$

where $a_1, a_2, \ldots, and\ a_n$ are the amount of NaCl or MHV-A59 captured in the impinger in filter-on experiments, and $b_1, b_2, \ldots, and\ b_m$ are the amount of NaCl or coronavirus captured in the impinger in filter-off experiments.

**Text S10. Electrospun Membranes Rather Than PP Fabrics Determining Filtration Performance for Aerosol Removal for the Electrospun Air Filters**

The PP fabrics that were used to support the electrospun membranes removed a limited amount of aerosols (0-30% aerosols based on NaCl aerosol filtration tests). We assume that the filtration efficiency of coronavirus aerosols was the same as that for NaCl aerosols, and thus at least 70% of coronaviruses in the aerosols passed through the PP fabric. In order to achieve an overall 99.1% removal of the coronavirus aerosols (an average filtration efficiency for $PVDF_{20}$), the electrospun layer had to remove at least 98.7% (= 1-(1-99.1%)/(1-30%)) of the coronaviruses in the aerosols. Therefore, the electrospun membranes rather than PP fabrics determining the filtration efficiency.

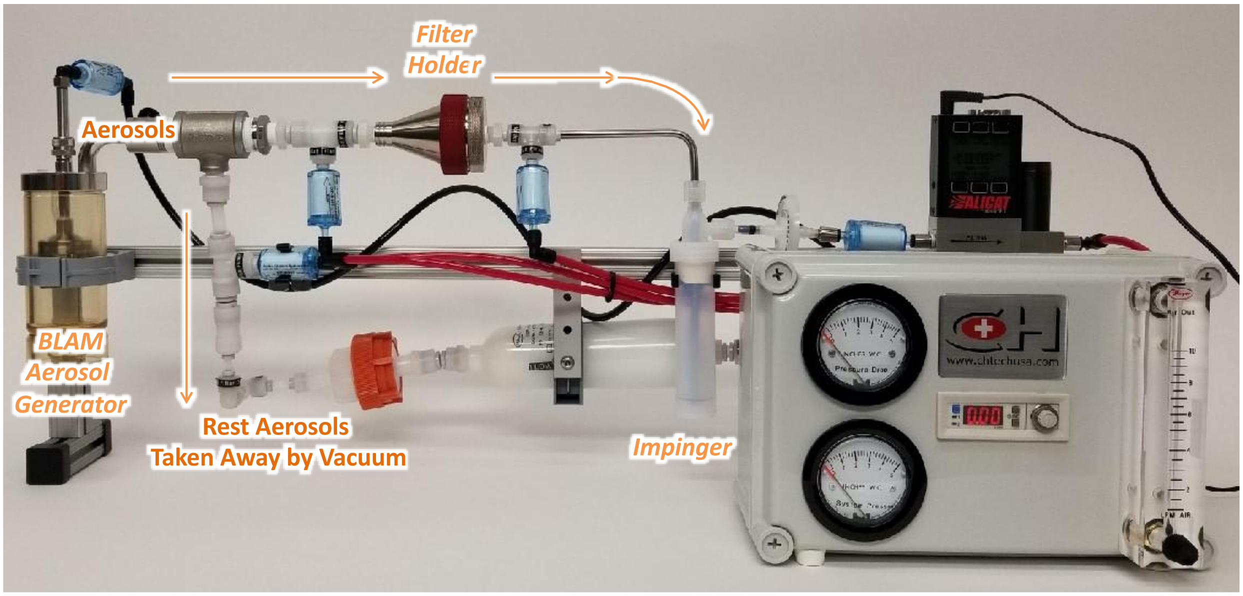


**Figure S1.** Scheme of the air filtration setup.

(a)

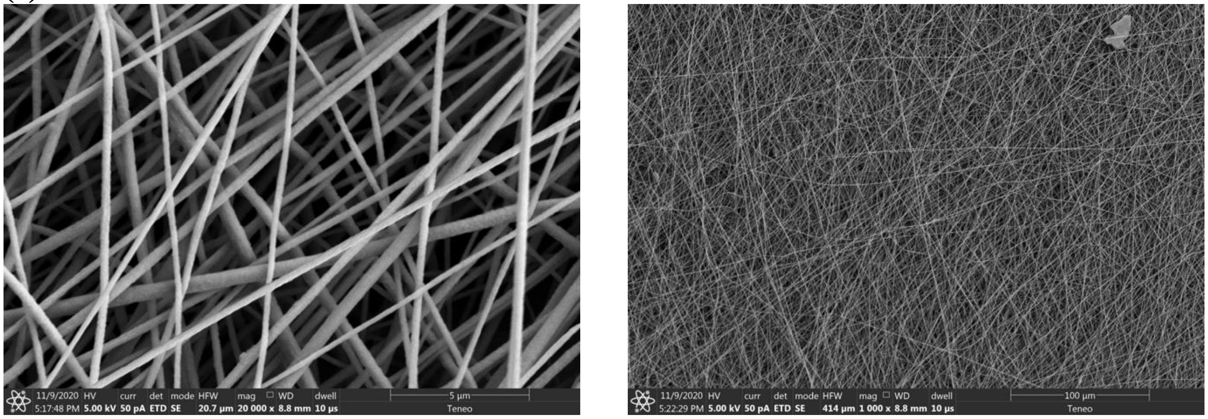
11/9/2020 HV curr det mode HFW mag WD dwell
5:17:48 PM 5.00 kV 50 pA ETD SE 20.7 µm 20 000 x 8.8 mm 10 µs
5 µm
Teneo
11/9/2020 HV curr det mode HFW mag WD dwell
5:22:29 PM 5.00 kV 50 pA ETD SE 414 µm 1 000 x 8.8 mm 10 µs
100 µm
Teneo

(b)

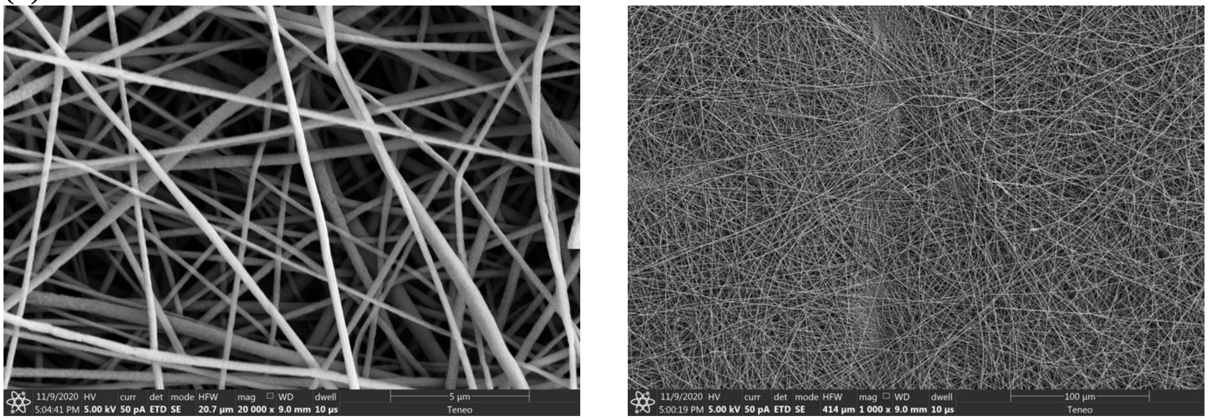
11/9/2020 HV curr det mode HFW mag WD dwell
5:04:41 PM 5.00 kV 50 pA ETD SE 20.7 µm 20 000 x 9.0 mm 10 µs
5 µm
Teneo
11/9/2020 HV curr det mode HFW mag WD dwell
5:00:19 PM 5.00 kV 50 pA ETD SE 414 µm 1 000 x 9.0 mm 10 µs
100 µm
Teneo

(c)

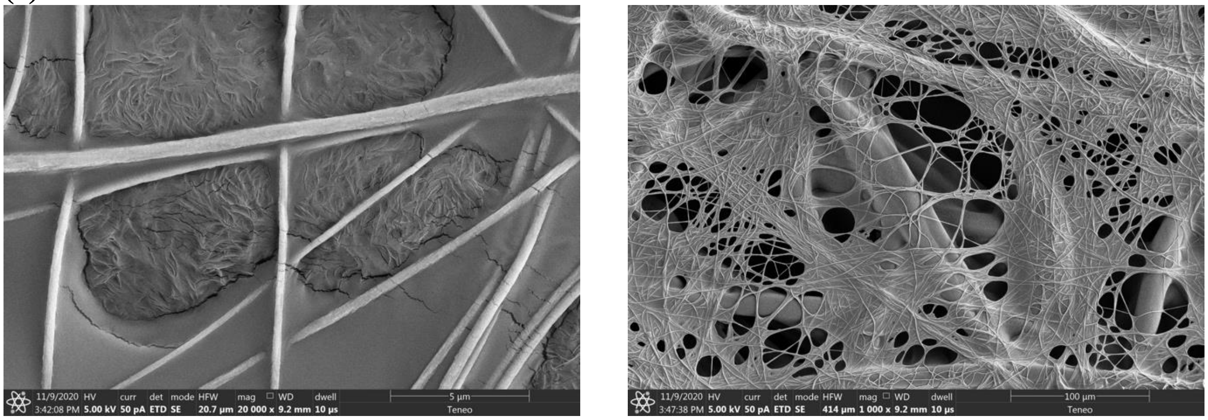
11/9/2020 HV curr det mode HFW mag WD dwell
3:42:08 PM 5.00 kV 50 pA ETD SE 20.7 µm 20 000 x 9.2 mm 10 µs
5 µm
Teneo
11/9/2020 HV curr det mode HFW mag WD dwell
3:47:38 PM 5.00 kV 50 pA ETD SE 414 µm 1 000 x 9.2 mm 10 µs
100 µm
Teneo

(d)

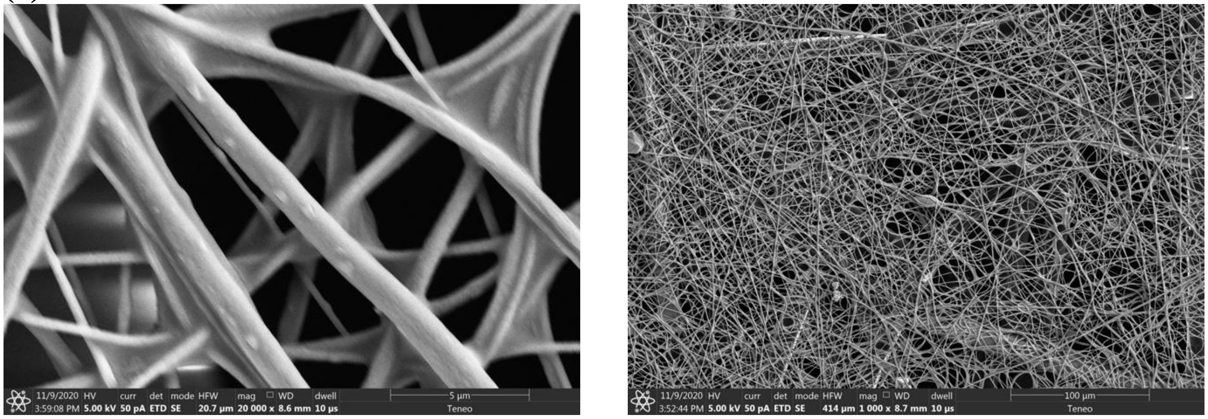

(e)

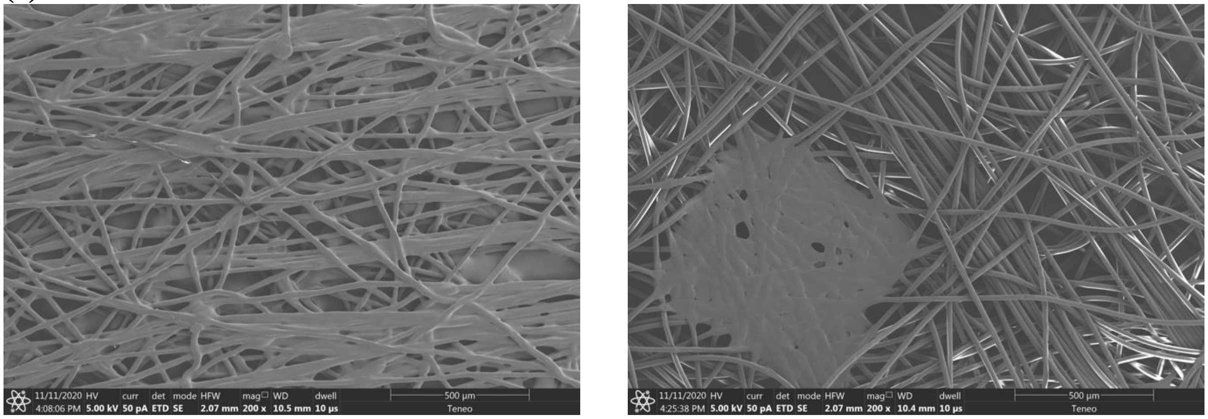

(f)

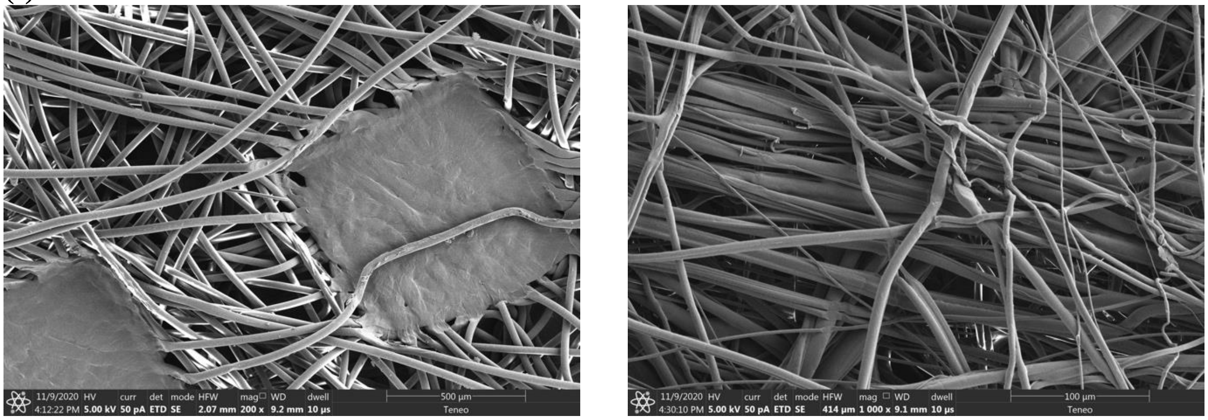

Outer layer Middle layer

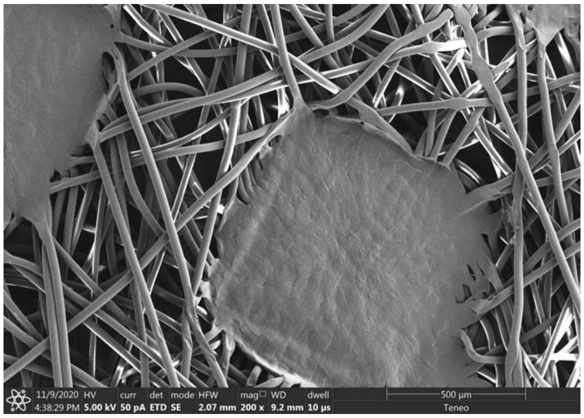

Inner layer

(g)

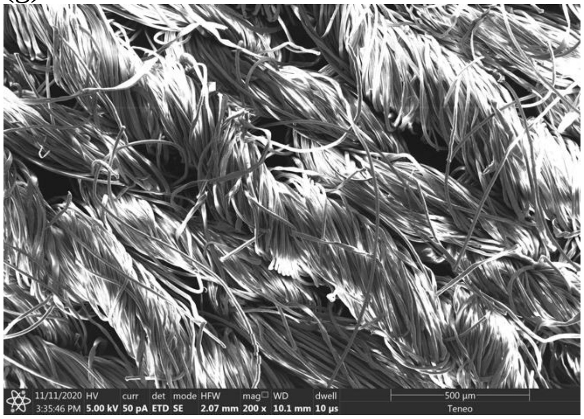

Outer layer

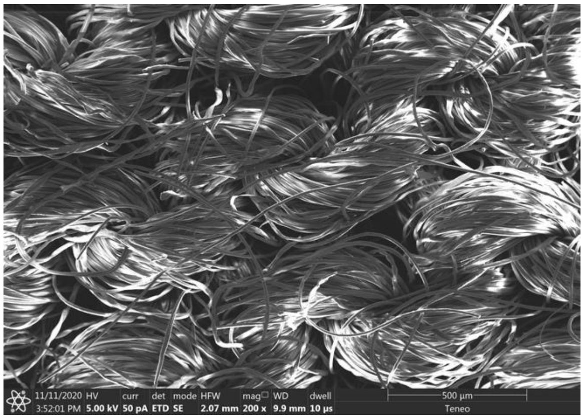

Middle layer

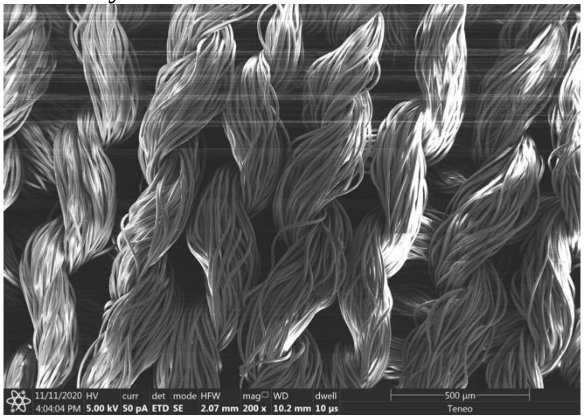

Inner layer

(h)

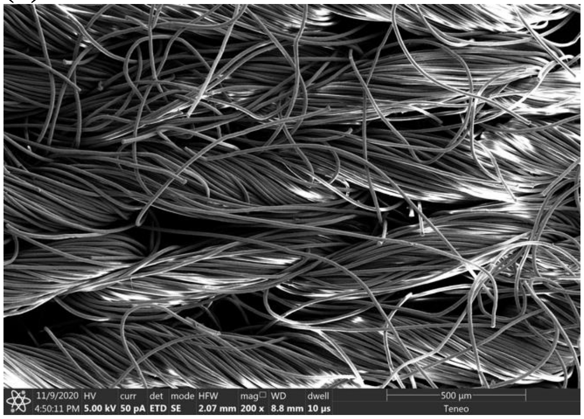


**Figure S2.** SEM of (a) $PVDF_{20}$, (b) $PVDF_{30}$, (c) $PVDF_{20}$/PEI, (d) $PVDF_{20}$/PVPA, (e) PP fabrics from VWR basic protection face mask, (f) surgical mask, (g) cotton mask, and (h) neck gaiter.

(a)

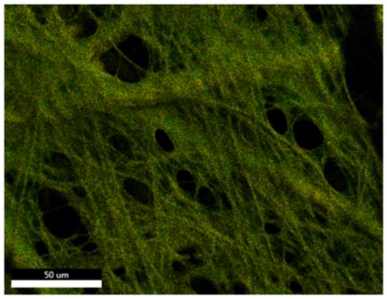


C and N

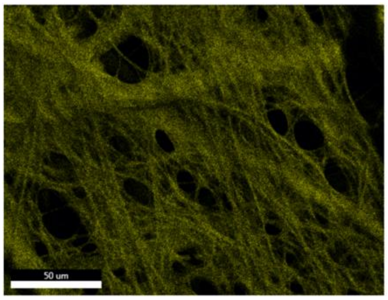


C

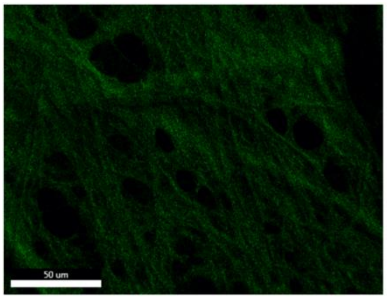


N

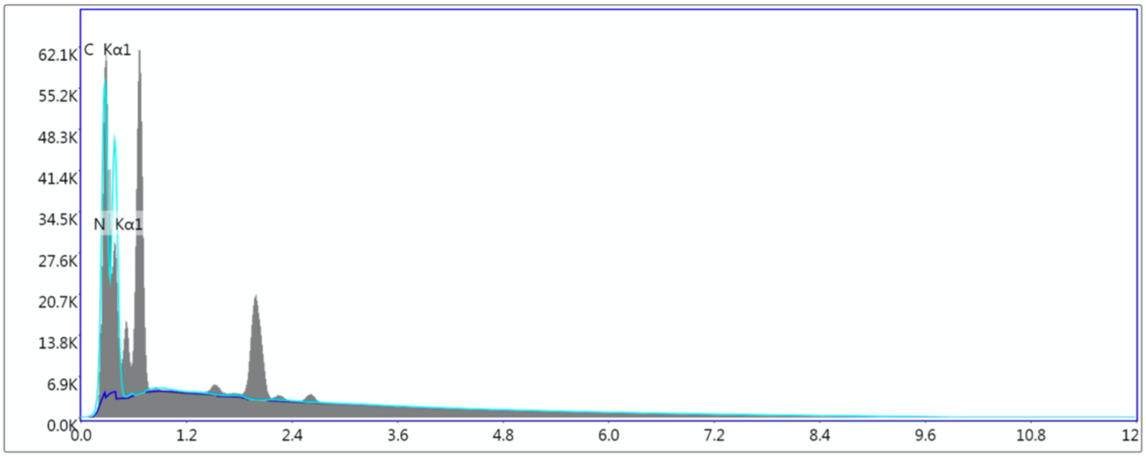


(b)

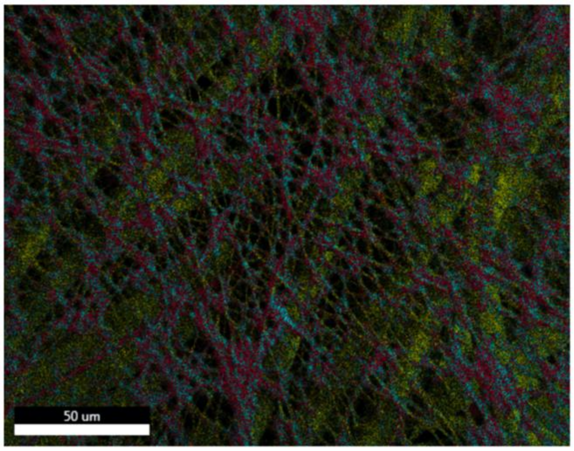


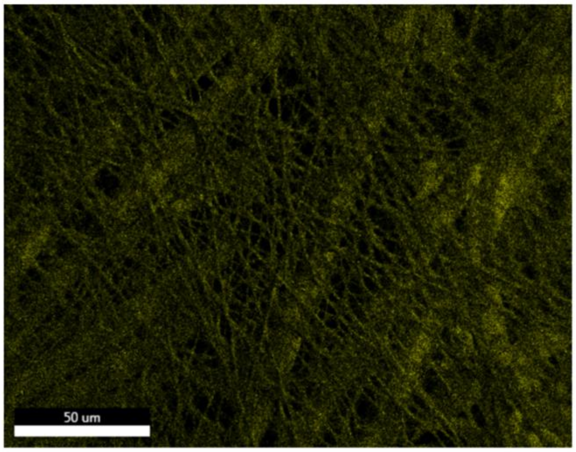


C, O, and P

C

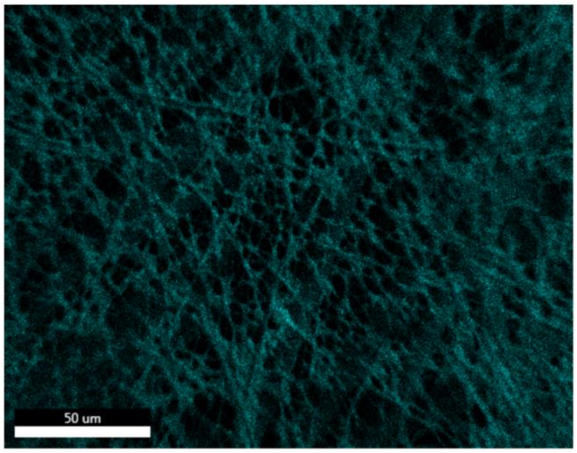


O

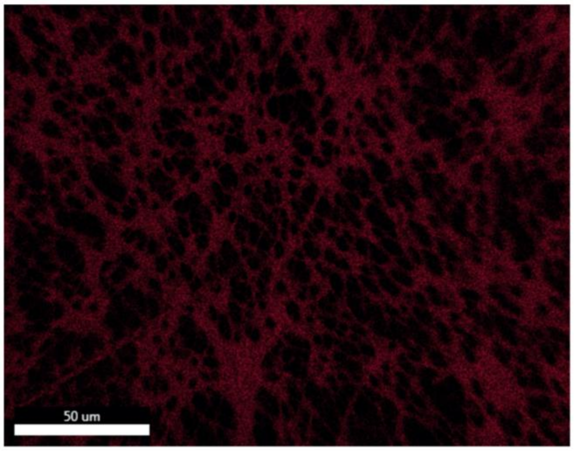


P

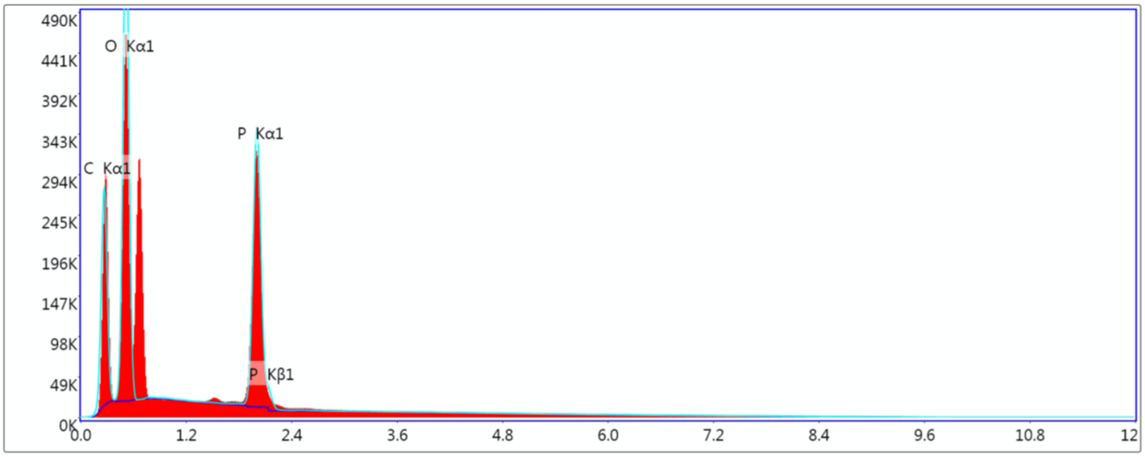


**Figure S3.** Elemental mapping of (a) $PVDF_{20}$/PEI and (b) $PVDF_{20}$/PVPA characterized by SEM-EDS. PEI has a unique element of N while PVPA has O and P compared to PVDF, so the presence of these elements indicates the successful coating of PEI or PVPA onto electrospun PVDF.

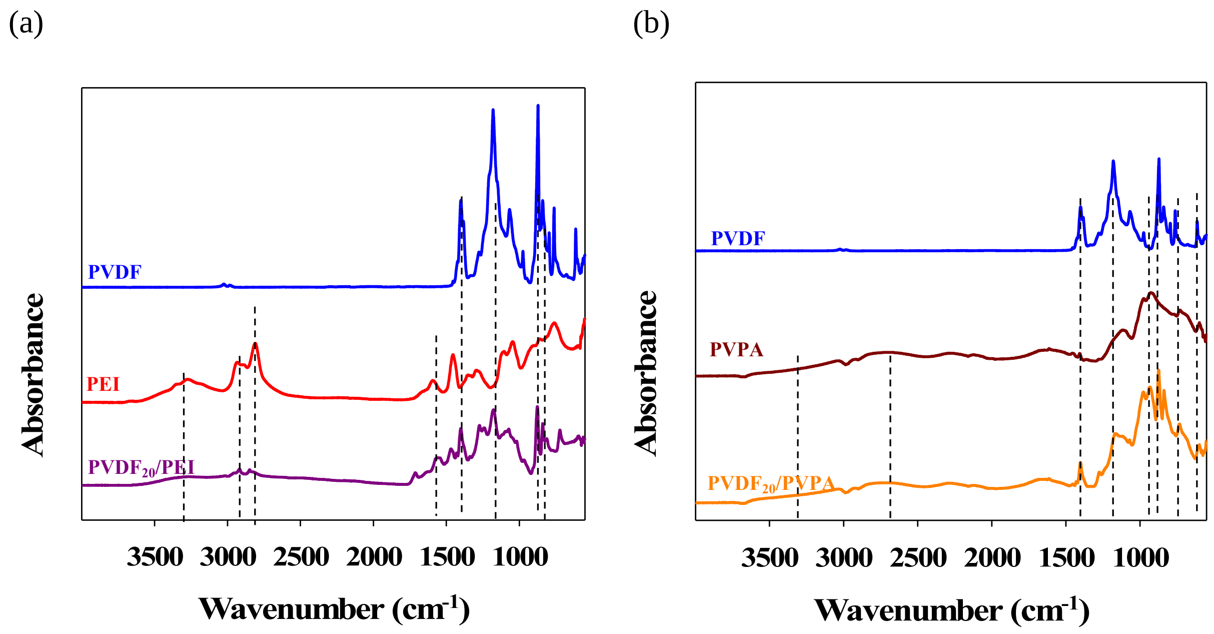


**Figure S4.** ATR-FTIR of (a) $PVDF_{20}$/PEI and (b) $PVDF_{20}$/PVPA. PVDF showed stretches at 1401, 1180, 1067, and 872 $cm^{-1}$; PEI showed stretches at 3272, 2933, 2809, 1593, 1455, 1293, 1046, and 760 $cm^{-1}$; and PVPA showed stretches at 3500-3000, 2900-2500, 1406, 1116, 927, and 599 $cm^{-1}$. Polyelectrolyte-coated electrospun filters clearly share the dominant features of both PVDF and the polyelectrolyte, indicating successful coating of PEI or PVPA.
